# Predicting refugee flows from Ukraine with an approach to Big (Crisis) Data: a new opportunity for refugee and humanitarian studies

**DOI:** 10.1101/2022.03.15.22272428

**Authors:** Tado Jurić

## Abstract

**Background:** This paper shows that Big Data and the so-called tools of digital demography, such as Google Trends (GT) and insights from social networks such as Instagram, Twitter and Facebook, can be useful for determining, estimating, and predicting the forced migration flows to the EU caused by the war in Ukraine.

**Objective:** The objective of this study was to test the usefulness of Google Trends indexes to predict further forced migration from Ukraine to the EU (mainly to Germany) and gain demographic insights from social networks into the age and gender structure of refugees.

**Methods:** The primary methodological concept of our approach is to monitor the digital trace of Internet searches in Ukrainian, Russian and English with the Google Trends analytical tool (trends.google.com). Initially, keywords were chosen that are most predictive, specific, and common enough to predict the forced migration from Ukraine. We requested the data before and during the war outbreak and divided the keyword frequency for each migration-related query to standardise the data. We compared this search frequency index with official statistics from UNHCR to prove the significations of results and correlations and test the model’s predictive potential. Since UNHCR does not yet have complete data on the demographic structure of refugees, to fill this gap, we used three other alternative Big Data sources: Facebook, Twitter and Instagram.

**Results:** All tested migration-related search queries about emigration planning from Ukraine show the positive linear association between Google index and data from official UNHCR statistics; R^2^ = 0.1211 for searches in Russian and R^2^ = 0.1831 for searches in Ukrainian. It is noticed that Ukrainians use the Russian language more often to search for terms than Ukrainian. Increase in migration-related search activities in Ukraine such as “граница” (Rus. border), кордону (Ukr. border); “Польща” (Poland); “Германия” (Rus. Germany), “Німеччина” (Ukr. Germany) and “Угорщина” and “Венгрия” (Hungary) correlate strongly with officially UNHCR data for externally displaced persons from Ukraine. All three languages show that the interest in Poland is the highest. When refugees arrive in nearby countries, the search for terms related to *Germany*, such as “crossing the border + Germany”, etc., is proliferating. This result confirms our hypothesis that one-third of all refugees will cross into Germany. According to Big Data insights, the estimate of the total number of expected refugees is to expect 5,4 Million refugees. The age group most represented is between 24 and 45 years (data for children are unavailable), and over 65% are women.

**Conclusion:** The increase in migration-related search queries is correlated with the rise in the number of refugees from Ukraine in the EU. Thus this method allows reliable forecasts. Understanding the consequences of forced migration from Ukraine is crucial to enabling UNHCR and governments to develop optimal humanitarian strategies and prepare for refugee reception and possible integration. The benefit of this method is reliable estimates and forecasting that can allow governments and UNHCR to prepare and better respond to the recent humanitarian crisis.

## Introduction

The Ukraine war that started on February 24 2022 destroyed civilian infrastructure and forced people to flee their homes to seek safety, protection, and assistance. According to UNHCR, more than a million refugees from Ukraine crossed borders into neighbouring countries in the first week since the war outbreak (2,5 Mil. in the second week), and many more are on the move both inside and outside the country. As the situation unfolds, an estimated 4 million people may flee Ukraine.^1^ The UNHCR estimated on February 27 that in two months, there would be 7.5 million internally displaced people in Ukraine. According to the UNHCR, 18 million people are affected by the conflict, and 12 million people will require health care.^2^

Nobody knows how long the war will last, what kind of conflict this will develop into, and the civilian toll. We know only that the sudden and massive flow of Ukrainians to Poland, Hungary, Slovakia, Romania and Moldova stretched UNHCR and humanitarian organisations to the limits of their capacities.

The crisis is just one of many during which having reliable data would have assisted UNHCR and UN agencies in preparing high-quality projections for emergencies. But such data are often either unavailable or only available with a considerable time lag, which renders them useless for operations and emergency preparedness.^3^ This study was created due to the need to predict migration flows of refugees from Ukraine to the EU. We present a descriptive analysis of Big Data sources, which could be helpful in determining, as well as for estimating and forecasting refuge emigration flows from Ukraine.

The expression “Big Data” has been spreading since 2011; the term is used in academia, industry and the media, but it is not even today precisely clear what it means. In this study, the term “Big Data” refers to the interdisciplinary method for deriving new understanding from massive aggregations of the information sampled through Google analytical tools Google Trends as well as social networks. We refer to the term Big (Crisis) Data as the subset of Big Data sources that have been applied in the humanitarian work of UNHCR.^4^

Since there has been a substantial increase in internet access compared to creating credible migration monitoring registration systems^5^, the development of statistical tools that combine traditional and new sources of information is likely to become an accepted approach to monitoring migration and refugee flows. In demography, researchers have begun to use non-traditional or alternative data (mobile phone records, social media use, satellite maps, and internet-based platforms) to understand migration and mobility in light of new methodological approaches.^6^ This study will show that the analytical tool Google Trends (GT) and insights from social media can give valuable complementary data in the field of migration and refuge studies and be useful in the current refugee crisis.

The structure of the paper is as follows: after briefly showing the results of relevant studies, we explain this study’s methods and show the limitations of this method. We then discuss refugee flows from Ukraine and reveals the results achieved via this approach. In the section on results, we show the correlation between the Google search index and the official UNHCR statistics, as well as discuss how to forecast migration flows with Google Trends and what insights about refugees can we gain from social media.

### Forced migration from Ukraine

Ukraine was one of 15 republics of the Soviet Union and became an independent state in 1991^7^; it had a population of approximately 44 million. Since the end of the Soviet Union, the country had been a country of origin, transit and arrival for forced migration, as well as a provider of “durable solutions” for ethnic Ukrainians returning to the country from which they had been exiled during the Soviet era.^8^ In the 1990s, the ethnic composition of Ukraine changed significantly. Many ethnic Russians left, and many ethnic Ukrainians returned, including Tatars originally from Crimea, whose returned population increased fivefold between 1989 and 2001. The suspension of negotiations of the EU association agreement by a pro-Russia government in 2013 led to the Maidan protests in Kyiv and a pro-European regime change the following year, rapidly followed by Russia’s occupation of Crimea (2014).^9^ 5.2 million people were affected by the conflict, and 1.6 million were displaced within and outside Ukrainian borders.^10^ As of 2019, there were 1.4 million internally displaced people in Ukraine. Most Ukrainians sought refuge in Russia, but many also went to EU countries.

In refugee, migration and humanitarian studies, little attention has been given to the continued refugee resettlement of Ukrainians. A larger body of work can be found in area studies.^11^ This includes research on the Ukrainian internally displaced people (IDP)^12^ in Crimea^13^ and Donbas^14^ and the humanitarian crisis.^15^

In the recent exodus from Ukraine, UNHCR estimates that over 4 million people could flee from Ukraine and seek protection and support across the region. ^16^ Our estimates obtained with Big Data show that there will be at least 5.4 million refugees. According to data from UNHCR, the speed of the exodus is already more extensive than the migration crisis of 2015, when 1.3 million asylum seekers from Syria, Iraq, Afghanistan and Africa, fleeing poverty and wars, entered Europe.^17^ “This is the fastest-moving refugee crisis in Europe since the end of the second world war.”^18^

Most Ukrainians (53%) fled in the first week to Poland, which has welcomed at 04.03.2022 about 756,303 people, followed by Hungary, with 157,004. The number of refugees is also high in Hungary, Romania, Slovakia and Moldova. But not everyone decides to stay in the countries they first arrive in from Ukraine. For example, 140,000 people who came to Romania during the first eight days of the war travelled to other countries, leaving about 60,000 in Romania, according to the UNHCR.^19^

**Table 1.**
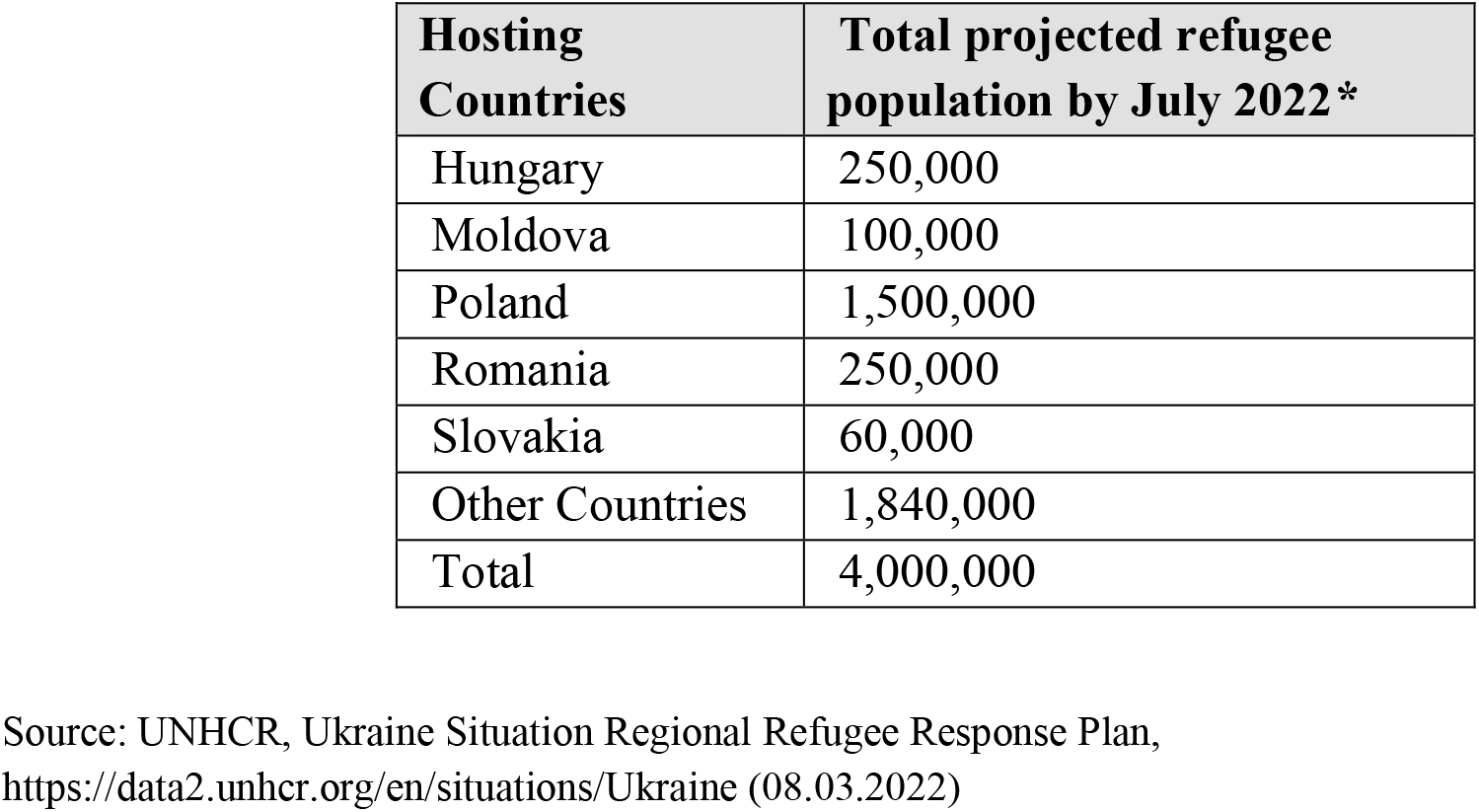
A total projected refugee from Ukraine by July 2022 (UNHCR)

| Hosting Countries | Total projected refugee population by July 2022* |
| --- | --- |
| Hungary | 250,000 |
| Moldova | 100,000 |
| Poland | 1,500,000 |
| Romania | 250,000 |
| Slovakia | 60,000 |
| Other Countries | 1,840,000 |
| Total | 4,000,000 |

Source: UNHCR, Ukraine Situation Regional Refugee Response Plan, https://data2.unhcr.org/en/situations/Ukraine (08.03.2022)

### Refuge welcome policy in the EU

In just one week, in the EU arrived so many refugees as in the entire so-called Yugoslav war.^20^ In the light of the current crisis, we believe that it is useful here to look back at the breakup of Yugoslavia and try to predict the course of the refugee crisis from lessons from the past. During the Homeland War in Croatia, 150,000 Croatians, or 3% of Croatia’s population, emigrated to Germany, while about 250,000 arrived from BiH. Of that, one third remained there forever, one third returned because their temporary stay (so-called Duldung) expired, while one third returned of their own free will.^21^

If we draw a parallel between the war in Ukraine and the Homeland War and assume that the same number of Ukrainians as former Croatians under the pressure of war will escape to Germany, we expect that at least 1,350,000 Ukrainians will emigrate to Germany. We assume that further migration waves will occur in the years to come when men are allowed to leave the country and emigrate to Germany due to family reunification and other reasons (for now, men between the ages of 18 and 60 cannot leave the country).

There are numerous differences in the welcome policy of Germany today and Germany in the 1990s. For entry to Germany during the Homeland war in Croatia, Croats asked the so-called letter of guarantee, i.e. proof that one of the relatives, etc., will guarantee the care of refugees. At the supranational level in the current crisis, the EU has triggered the adoption of the EU temporary protection directive for the first time. The directive grants immediate protection to Ukrainians fleeing to war for three years.^22^ Ukrainians don’t require Schengen visas to enter the EU, and there is no other restriction. The EU admits Ukrainians without valid passports if they pass an individual assessment conducted by border officials. Moreover, Germany openly encourages the immigration of Ukrainians. Namely, Ukrainian nationals will be given the right to live and work in the European Union for up to three years without claiming asylum.^23^

What has changed from the 1990s to today in Germany? In short, the demographic picture. Today, due to the demographic crisis in its own country, the lack of labour force of all profiles and young people in general, Germany needs to import about 400,000 employees every year to maintain its pension and health care system, according to a study by Bertelsmann Stiftung.^24^ According to projections, the potential labour force in Germany will decrease by 16.2 million workers between 2012 and 2050 for purely demographic reasons.^25^ The German generations with the highest birth rates will have left working life around 2035. According to model calculations, the net migration with countries of the EU will soon drop significantly from the current number to slightly below 300,000.^26^ For the next 36 years, an average of between 276,000 and 491,000 people would have to immigrate from third countries every year to Germany for the labour force potential to remain constant.^27^

Our primary hypothesis is that up to one-third of all refugees will cross into Germany. Surely, no refugee from Ukraine will be forced to stay in Germany, as neither are Croats during the Homeland War, but the combination of fear, a better standard and open hospitality will influence this decision.

The German Trade Union Confederation (DGB) and the German Employers’ Union (BDA) issued a joint statement calling on the German government to urgently remove “legal and bureaucratic” obstacles to the rapid integration of Ukrainian refugees into the German labour market. “Companies and works councils are ready to take their share of responsibility in order to take over these people as soon as possible, train or retrain them and thus integrate them into the labour market,” the joint statement said.^28^ Furthermore, the German Railways (Deutsche Bahn), three days after the start of the war, already on February 27 put at its disposal trains for free transport of all Ukrainians from Poland to Germany.

Some authors state that refugees from Syria and Africa have received worse treatment than refugees from Ukraine.^29^ In the 2015 European migrant crisis^30^, when 1.3 million people came to the continent to request asylum^31^, the welcome policy was completely different - for example, many EU members have lifted the razor wire on their borders. In March 2016, Turkey agreed to close its border to the EU in exchange for money and diplomatic favours, which effectively stopped the further passage of refugees through Eastern and South-eastern Europe.^32^

### Innovative data sources in the refuge and migration studies

Human migration within Europe is difficult to measure due to the lack of efficient statistical systems in numerous EU countries.^33^ Numerous researches have shown^34^ that obtaining timely evidence-based information on potential migratory movements is of the utmost importance to develop early-warning strategies and set up the necessary reception mechanisms in transit and receiving countries.

Traditional data sources, based either on surveys or registers, generally fail to quickly provide statistical information on refugee flows and do not facilitate the accurate short-term anticipation of these flows.^35^ This limitation is one reason underlying the development of new methods based on alternative sources, so-called Big Data.^36^

The potential of Big Data to facilitate early warning of crises lies in its ability to provide granular, almost real-time information in locations where there are few other data sources. According to UNHCR, some Big Data sources can have the power to fill some of the information gaps left by conventional data acquisition channels, especially during crises.^37^ According to Sales (2021), there are three main types of Big Data sources that are highly relevant to complement traditional migration and refugee data sources: communication services (calls records and texts messages), geo-localised activity as well as Internet-based exchanges (social media activity, online searching preferences, and online money transfers).^38^

The main advantages of this approach for refugee studies are that those data are easily collected and generated in real-time, they are incredibly robust, and they provide a profound insight into migrants’ or refugees’ opinions.^39^ This data can be used to gather insights into what was going on in the user’s mind through a non-invasive manner (see section Restrictions about ethical concerns).^40^ Moreover, digital traces provide documentation of both movement and activities, which can help researchers bypass possible sources of error in survey data, such as inability to recall, bias, and the like.^41^ Finally, digital traces can provide access to groups that are difficult to reach or are generally underrepresented by traditional research techniques. ^42^

According to Jurić ^43^, and Choi and Varian ^44^, Google Trends data have been used in various types of research: US unemployment, flu outbreak, predicting consumer behaviour, predicting inflation rates, predicting the housing market, predicting stock market changes, modelling tourism demand, etc. All the research results showed that GT analytical tools could reveal valuable insights about intentions.^45^

Several case studies have demonstrated that the inclusion of Big Data sources significantly improves the power and accuracy of predictive models of refugee flows.^46^ A study by Singh et al. (2019) on internally displaced persons (IDPs) movements between provinces in Iraq shows that a mix of social media data and traditional register data improves the predictive quality compared to predictions based on register data alone.^47^ Therefore, Big Data hold huge potential for humanitarian organisations to improve their early warning systems and generate better contingency planning figures. ^48^ Although previous research has established that digital data can be employed to study migration, there are still significant methodological issues and scepticism regarding the feasibility of using alternative data sources.^49^

In 2014 the UN conducted the first research on the use of Big Data for demographic research, with its report released in 2018.^50^ These data were shown to provide useful insights into the quantitative and qualitative characteristics of international and other migrations. After the UN confirmed the relevance of these data, explorations have been carried out on social networks^51^, and several studies have used Big Data sources to analyse migration-related phenomena directly.^52^

The European Commission concluded similarly to the UN that a) Big Data sources do not replace traditional data sources but can complement them, b) they can still be used to assess trends.^53^ Furthermore, it has been established that both kinds of data sources can complement each other.^54^ “In addition to being extremely robust, these data are easily collected, are generated in real-time, and provide significant insights into the opinions of individuals.”^55^

According to Gabrilovich, would-be migrants often use online searching to get answers about the country they plan to emigrate to^56^, and according to Wanner, Google is the first source of information for most users planning to relocate.^57^ Several studies have used immense data sources to analyse migration-related phenomena directly. The first successful analysis of this type of data was in 2009, and the first study in the field of migration examined during the 2015 Migration Crisis searches for particular terms in Arabic in Turkey and Germany according to selected terms such as “Greece” or “Germany”. ^58^ A study by the PEW - research centre showed that digital prints left by internet searches could provide insight into the movement of migrants. Namely, during their travels in 2015 and 2016, many migrants used smartphones that provided access to information and maps and travel tips via social media. It was then unequivocally proven that these indicators could be used to predict migration. ^59^ Compared to approaches using social media, the advantage of Google Trends is that limitations related to fake accounts are not prevalent.^60^

Undoubtedly, the Internet penetration rate is for this method significant. When it comes to the use of Internet services, Ukrainians are generally comparable to the EU average (with a slightly lower share).^61^ Migrants and refugees are nowadays increasingly using mobile devices and digital interactions on social media to communicate with relatives and send them news and pictures. Migrants use smartphones to facilitate their movements across borders, stay up to date with weather predictions, get last-minute information in transit and destination countries, or send and receive international remittances.^62^ Moreover, it is proven that the refugees are more interested in information from the Internet than the average and that they generally use smartphones during migration.^63^

## Methodological concept

The primary methodological concept of our approach is to monitor migration-related searches with the analytical tool Google Trends (GT) during the Ukraine war crisis.^64^ This tool shows the popularity of a specific term and shows if a trend is rising or falling. GT does not provide information on the actual number of keyword searches - instead, it standardises search volume on a scale of 0 to 100 over the period being examined, with higher values indicating the time when the search volume was greatest, allowing for verifiable metrics.^65^ In previous studies, it was important limitations that each of these searches was conducted for its reason and did not answer researchers’ questions, so “googling” the term “Germany” was not necessarily an implication that someone wants to move to Germany, but may be interested in tourist information or just looking for the German Bundesliga.^66^ However, in this crisis, it is quite certain that no one is interested in the Bundesliga when googling the word Germany. However, it is essential to pay attention to the overall context by interpreting the results.

The Google search Index cannot estimate the exact number of searches, so with the help of this tool, the exact number of emigrants cannot be calculated, but the increase of the trend can be noticed very precisely.^67^ This method we have already tested in our previous studies with quite good predictive indicators.^68^ The justification of using the GT method for assessment of refugee flows is that mismatch between intention and actual behaviour, which is one of the most significant restrictions of this approach, is in the case of refugees from Ukraine not to such an extent represented as in the case of predicting migrations in peacetime.^69^

For this method, it is essential to establish the time flow from the expressed intention to the realisation of migration. We hypothesise that an increase in the number of Google searches will, following a delay of about one to three days, translate to a rise in the number of refugees. According to Curry et al., trigger events happen immediately before migration, usually within a time frame of 1-2 days before emigration occurs, and are events that the individual perceives as threatening their integrity.^70^

To standardise the data for Google Trends, we extracted data from February 24 2021, to February 24 2022. We then divided the keyword frequency for each migration-related word, which gave us a search frequency index that we then compared with official statistics using a linear regression method.^71^ To estimate the model, linear regression was used to measure the correlation between the number of searches (x) and the number of moves (y) evidenced by the official UNHCR statistics.

Initially, keywords were chosen by brainstorming possible words that we believed to be predictive, specific, and common enough to forecast refugees flows. After the significance screen, we selected followed keywords and topics.

**Table 2.**
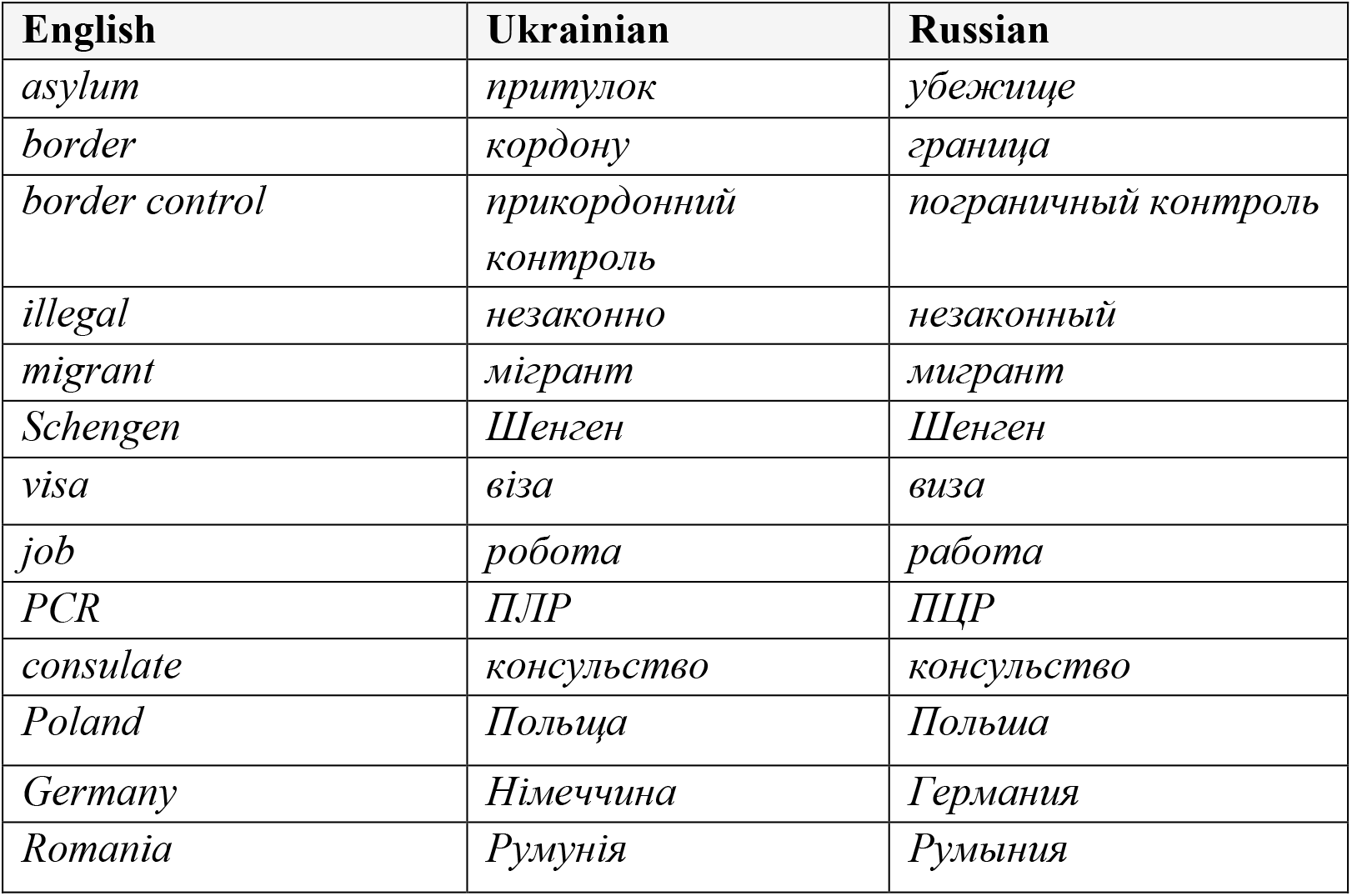
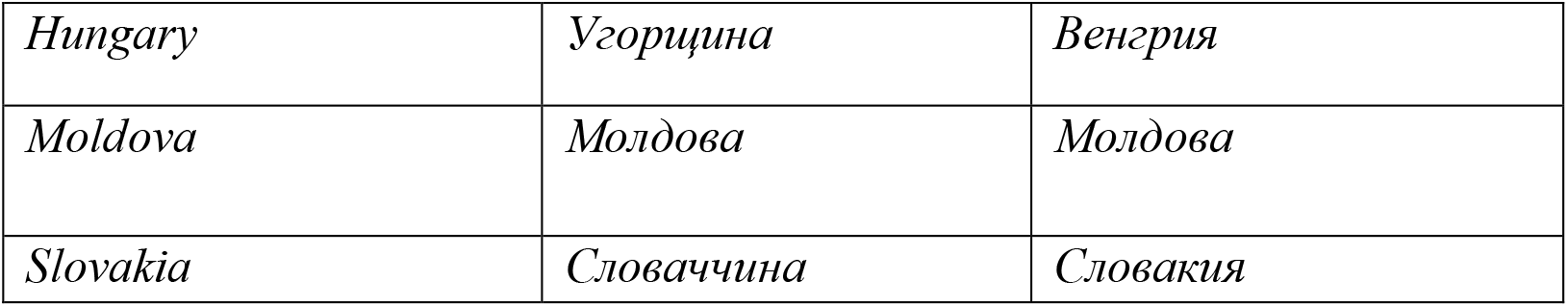
List of Keywords.

It is to note that we have detected that Ukrainians use the Russian language more often to search for terms than Ukrainian. Of the foreign languages, English is most often used.

### Limitations of the methodological concept

Like all data sources, Big Data also come with limitations. According to UNHCR, issues with Big Data mostly come from three sources: bias, inaccuracy, and low scalability.^72^ GT has some specific limitations when applied in the context of forced displacement. Namely, many displaced people do have not the time to conduct detailed searches on the Internet for a potential host country.^73^ But the biggest weakness of GT data, which is biased due to limited access to the Internet, is in the case of refugees from Ukraine not represented. As already mentioned, it is proved that refugees are ready to allocate the most resources for smartphones, i.e. connection with relatives and friends, but also the Internet.

Although penetration rates of the Internet are increasing globally, they still do not reach 100% of the Ukrainian population. Rural communities, women, children, and the elderly are underrepresented population segments and risk becoming invisible in Big Data sources.^74^ On the other hand, there are numerous indications that potential migrants and refugees largely gather or refine information through Google searches before emigrating.^75^ According to a Europol report (2018), there has been an exponential growth in the use of the Internet and social media by refugees and migrants arriving in Europe in the last few years.^76^ Even the UNHCR has concluded that smartphones, the Internet and social media are now a key tool for migrants, who spend up to a third of their total budget on staying connected.^77^

Although the data obtained with GT are robust data with large samples, which provide information qualitatively different from what can be obtained from the official databases, they are not representative of the observed population.^78^ A significant restriction is that GT does not provide data on which population was sampled or how it was structured.^79^ A problem also exists in the researchers’ education who must be skilled in computational methods, be transparent about their methods to ensure repeatability, and be accustomed to the interdisciplinary environment.^80^

We are also aware that refugees may use other methods to gather information on living and working conditions in Poland and Germany, such as the accounts of friends or family members who have already immigrated to these countries. A further aspect is the availability of humanitarian help in a potential host country, a network within an existing diaspora in the likely host country, and available information on flight routes and corridors from earlier migrants.^81^

Regarding restrictions in social media data, the problem with Facebook and Instagram data lies in the possibility that users have multiple unconnected accounts, which can produce data distortion. A certain number of users might be our target group but use Facebook in another language (instead of Ukrainian, Russian or English). Furthermore, a problem arises with the withdrawal of data from Facebook and Instagram because it is possible to get data only for the current day. The researcher, therefore, must collect data for his own archive every day.

Twitter data is only effective if the user uses the geotagging option, typically using only 3% of users. Furthermore, the basic idea of monitoring the moves of refugees from country to country through geotagged messages usually requires a certain longer period, which in our study, due to actuality, could not be done.

Regarding ethical issues, since the vast majority of Big Data is generated automatically by mobile and Internet users without informed consent and knowledge for what purpose those data are collected, its use may lead to violations on the grounds of privacy and data protection.^82^ This situation also raises concerns about public-private cooperation because governments may become dependent on those Big Data collection.^83^

## Results

### Use of the Google Trends analytical tool to forecast refugee flows from Ukraine

Searching for queries in Ukrainian, Russian and English from Ukraine “border crossing”, “кордону” (Ukr. border), “граница” (Rus. border) from December 7, 2021, to March 3 2022 (Figure 2) shows an upward trend since the outbreak of war. This is a strong indication that more and more Ukrainian citizens will emigrate, i.e. flee from Ukraine.

**Figure 1:**
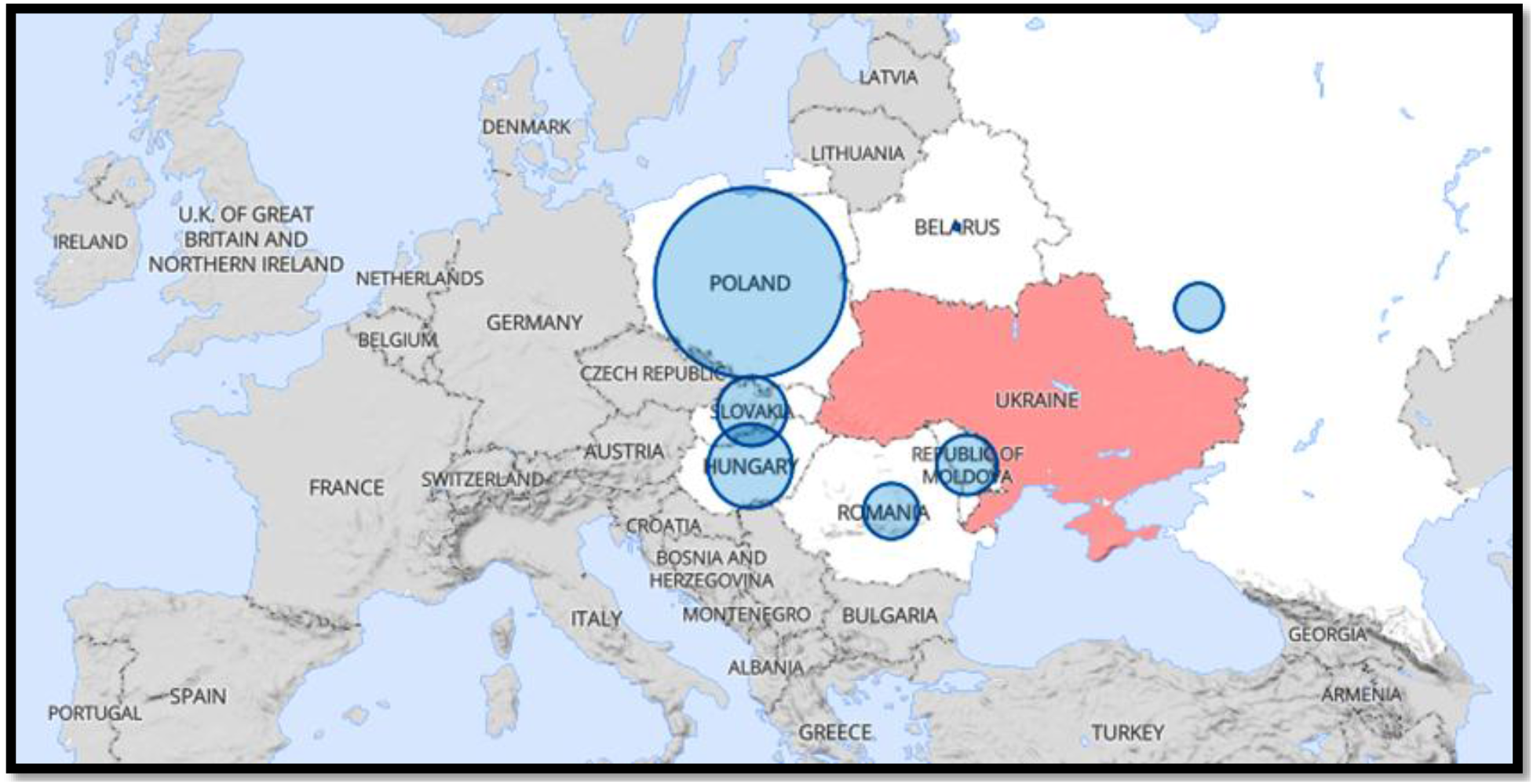
Refugee from Ukraine (UNHCR) Source: UNHCR, https://data2.unhcr.org/en/situations/Ukraine (08.03.2022)

**Figure 2.**
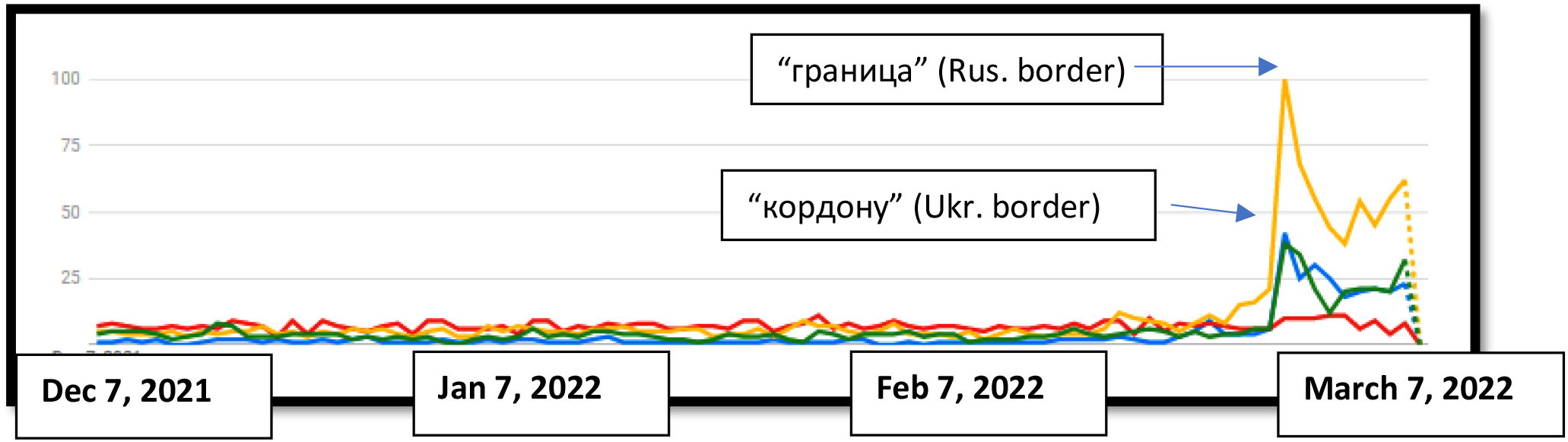
Search queries in Ukrainian, Russian and English from Ukraine “border crossing”, “кордону” (Ukr. border), “граница” (Rus. border) (December 7 2021 – March 3 2022)

A further indication is searching for terms related to registration in Poland, Hungary and Germany.

Figure 3 shows that the fastest-growing Google search terms in Ukraine (December 7 to March 7), except mentioned term “border”, are Western Union, asylum, refugee and Schengen.

**Figure 3.**
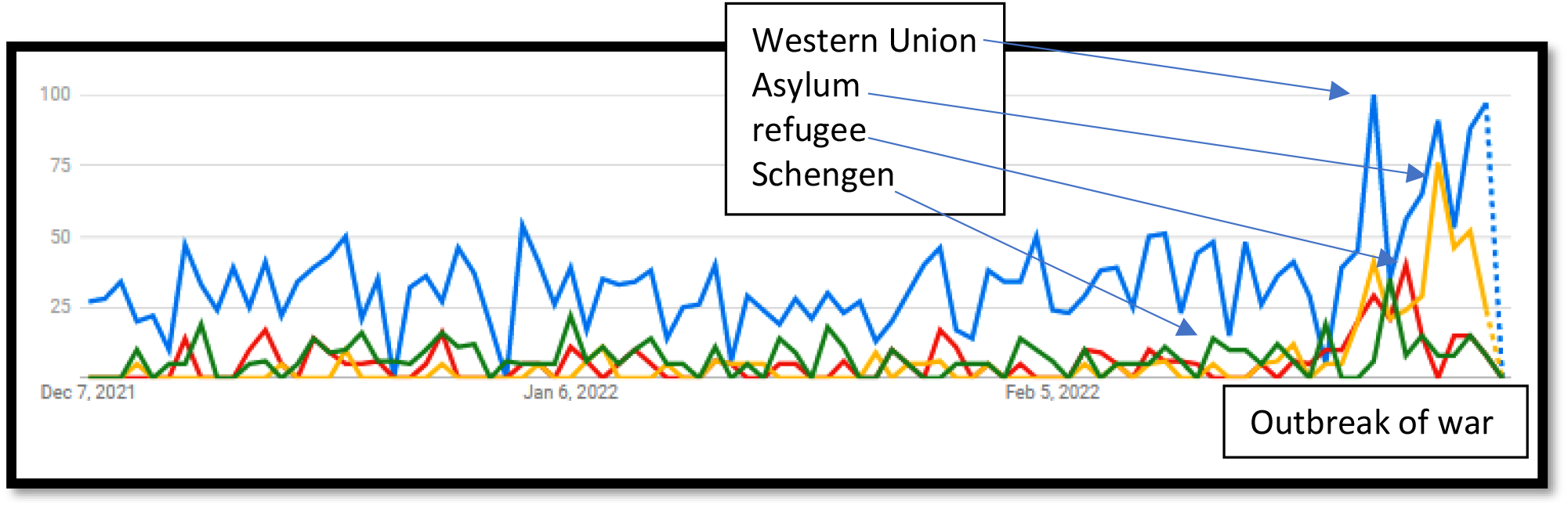
Fastest growing Google search terms in Ukraine: *Western Union, Asylum, refugee, Schengen* (December 7 to March 7)

When looking at the showed interest for emigration countries, we notice that the internet traces correspond to the official data of the UNHCR. Namely, the GT, just like the UNHCR, shows that the interest is focused primarily on Poland, Hungary, Slovakia and Germany (UNHCR still does not confirm information about Germany, March 11 2022).

Figure 4 shows that the most searched countries from Ukraine in Ukrainian are Poland and Hungary. As already mentioned, the citizens of Ukraine use Russian during Internet searches more than Ukrainian. This could be explained that the citizens of Ukraine probably expect more information in Russian, which is known as a world language, but also that the vast majority of Ukrainians learn and use Russian.

**Figure 4.**
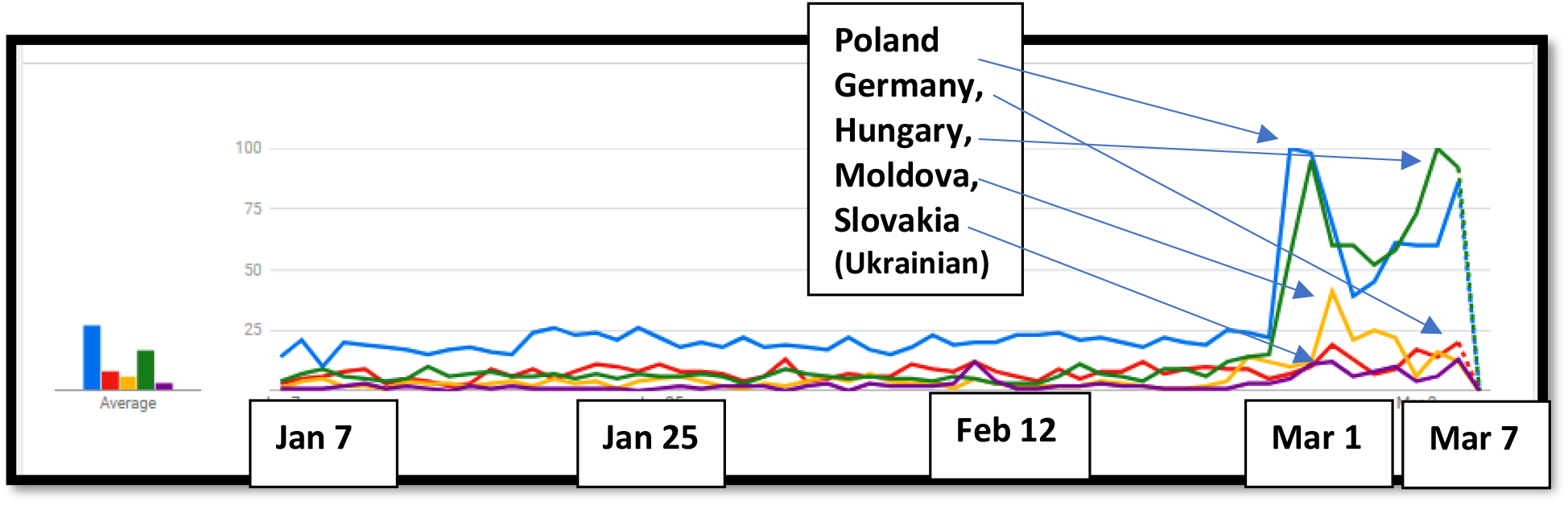
Search queries in Ukrainian from Ukraine “Польща”, “Німеччина” “Угорщина”, “Молдова” „Словаччина” (Poland, Germany, Hungary, Moldova, Slovakia) (January 7 2021 – March 3 2022)

Figure 5 shows that the most searched countries in Ukraine in Russian are Poland and Moldova. The following Figure 6 shows that *Poland* and *Romania* are the most searched countries in English.

**Figure 5.**
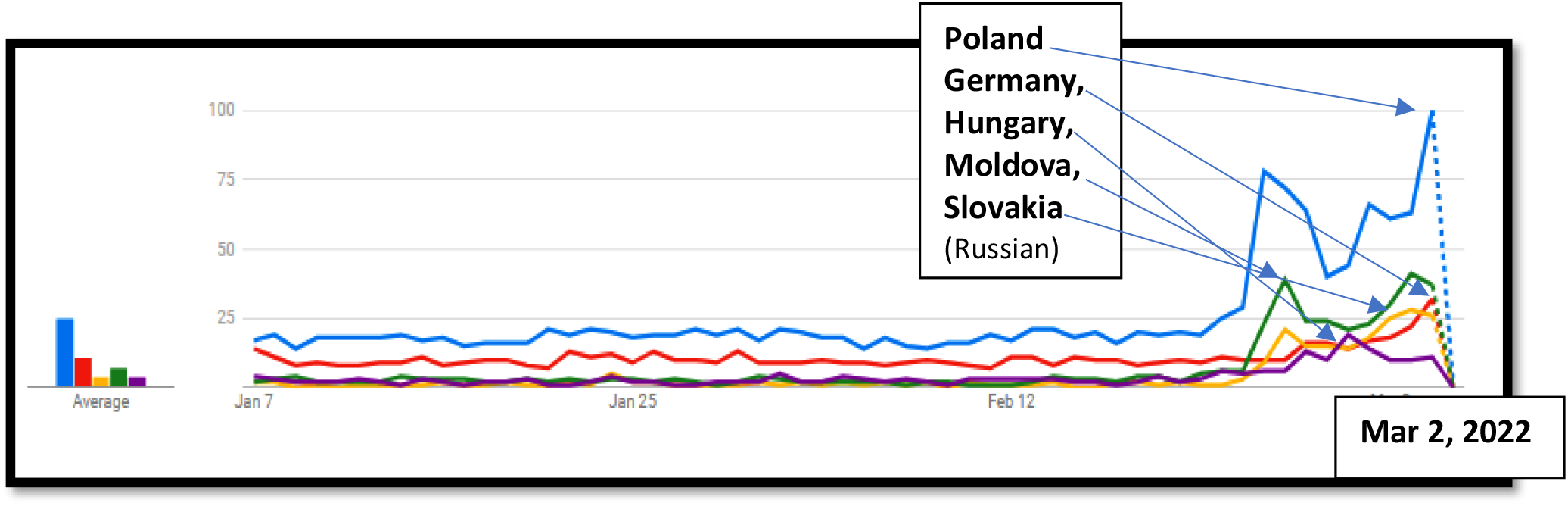
Search queries in Russian from Ukraine “Польша”, “Германия”, “Румыния”, “Венгрия”, “Молдова” (Poland, Germany, Hungary, Moldova, Slovakia) (January 7 2021 – March 3 2022)

**Figure 6.**
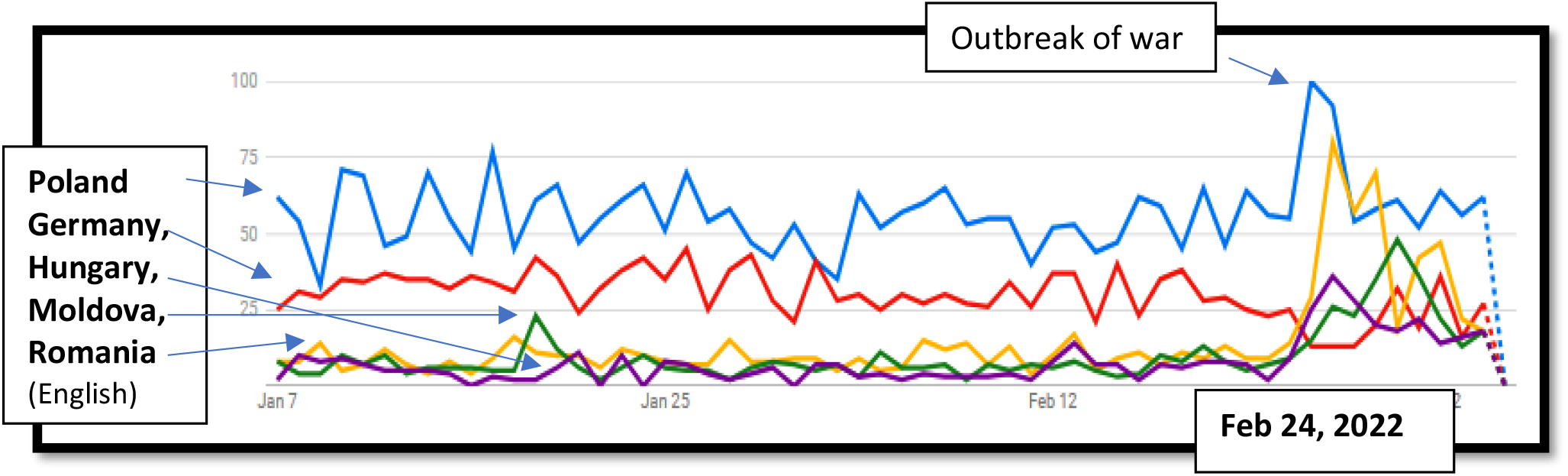
Search queries in English *Poland, Germany, Hungary, Moldova, Slovakia* from Ukraine (January 7 2021 – March 3 2022)

Figures 4, 5 and 6 show the increase in Internet searches for all neighbouring countries in Russian, Ukrainian and English; all three languages shows that the search interest for Poland is the highest. Other languages, including German, are not significantly represented.

Our basic hypothesis is that one-third of all refugees from Ukraine will emigrate further to Germany after arriving in the original EU countries. As we stated in the introduction, this assumption is based on the movement of refugees during the Homeland War in Croatia, when it turned out that most Croatian citizens, after fleeing to nearby countries, Hungary and Slovenia, continued to Germany. Undoubtedly, the economic power of the receiving country and the assessment of when the war could stop also play an important role in this prediction. The data we received with the GT application undoubtedly show a high increase in interest in Germany, but also a set of inquiries related to the so-called “German way of life”, jobs opportunities, children’s enrollment in school and other indicators that undoubtedly indicate the intention of refugees to stay longer in Germany.

Further confirmation of this hypothesis is the increase in specific searches that undoubtedly reveal the intention to move or flee to Germany. One of the most common searches is just focused on the question “Германия принимает беженцев из Украины” (Does Germany accepts refugees from Ukraine?).

This specific search also correlates with the regions most affected by the war, such as Donetsk, Luhansk, Kharkiv, Odesa and Kyiv.

As time goes on, refugees usually realise that the war and its consequences will, unfortunately, last longer than they expected. When they notice this, they continue their emigration to a country that offers the highest economic and financial security. In this case, as in the case of the Homeland War in Croatia, this is Germany. This assumption confirms the high growth of interest in Poland for the search query “Germany”.

The most frequent search in Poland since the outbreak of the Russian-Ukrainian war is “Border crossing+Germany”.

According to the German Federal Ministry of the Interior, 37,786 war refugees from Ukraine were registered in Germany by midday on March 6, 2022.^84^ The search for terms related to crossing the border into Germany in Poland is growing precisely in the regions located near Ukraine.

According to Düvell, there are currently up to 24 million people affected by the war. Half of the population was displaced in the Crimea-Donbass conflict. According to the UNHCR, 2.3 million citizens were displaced in Ukraine during the Crimean crisis. Of that, 1.5 million escaped from Crimea (800,000 were of Russian ethnicity).^85^ If half of the affected population flees like the Crimean occupation in 2014, there will be up to 12 million Ukrainian refugees.^86^ That this hypothesis could be correct, we see when we compare the search query “Германия принимает беженцев из Украины” (Does Germany accepts refugees from Ukraine) during the Annexation of Crimea in 2014 with the current crisis.

Figure 13 shows that during the peak of the Crimean crisis in 2014, the search index for query “Германия принимает беженцев из Украины” (Germany accepts refugees from Ukraine) was 4, and in 2022 it is 12, that is, the interest in fleeing the country is now three times higher.

**Figure 7.**
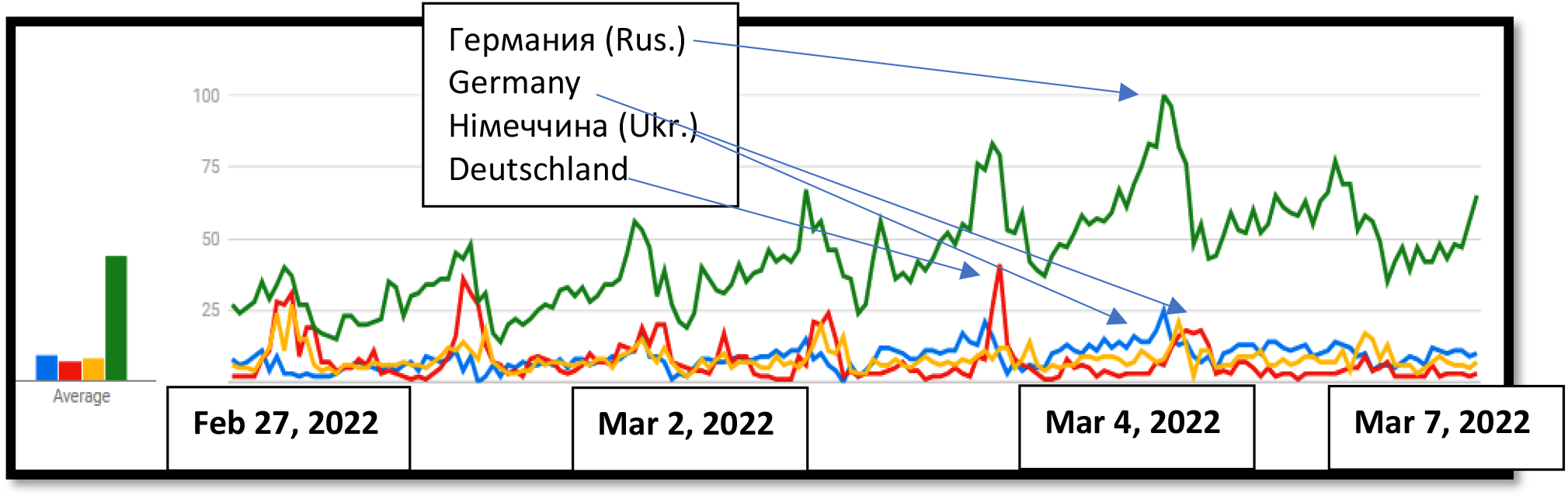
Interest in the search term “Germany” in Ukraine in English, German, Russian and Ukrainian since the outbreak of war (February 28 to Mar 06.2022)

**Figure 8.**
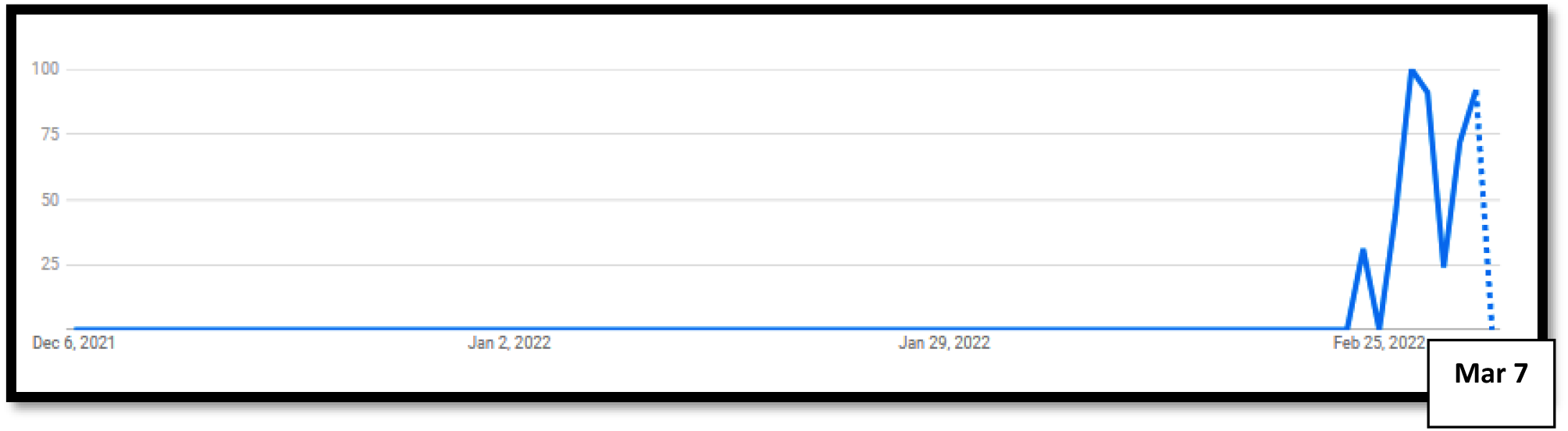
Increase in search query “Германия принимает беженцев из Украины” (Does Germany accepts refugees from Ukraine) (Dec 6 2021 to Mar 08 2022)

**Figure 9.**
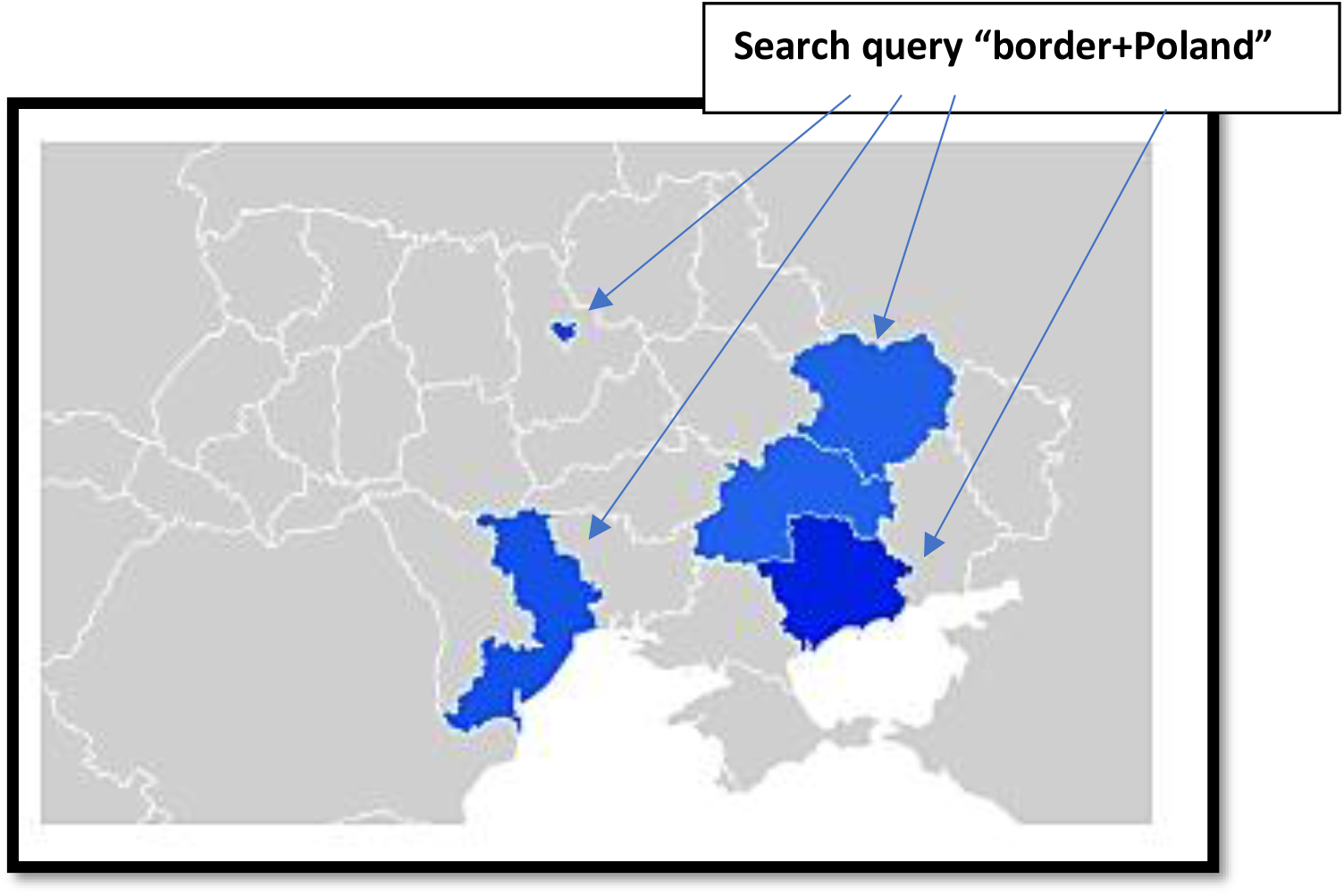
Correlation of Google search “border+Poland” with the regions in Ukraine most affected by the war, such as Donetsk, Lugansk, Harkiv, Odesa and Kyiv (March 6 2022)

**Figure 10.**
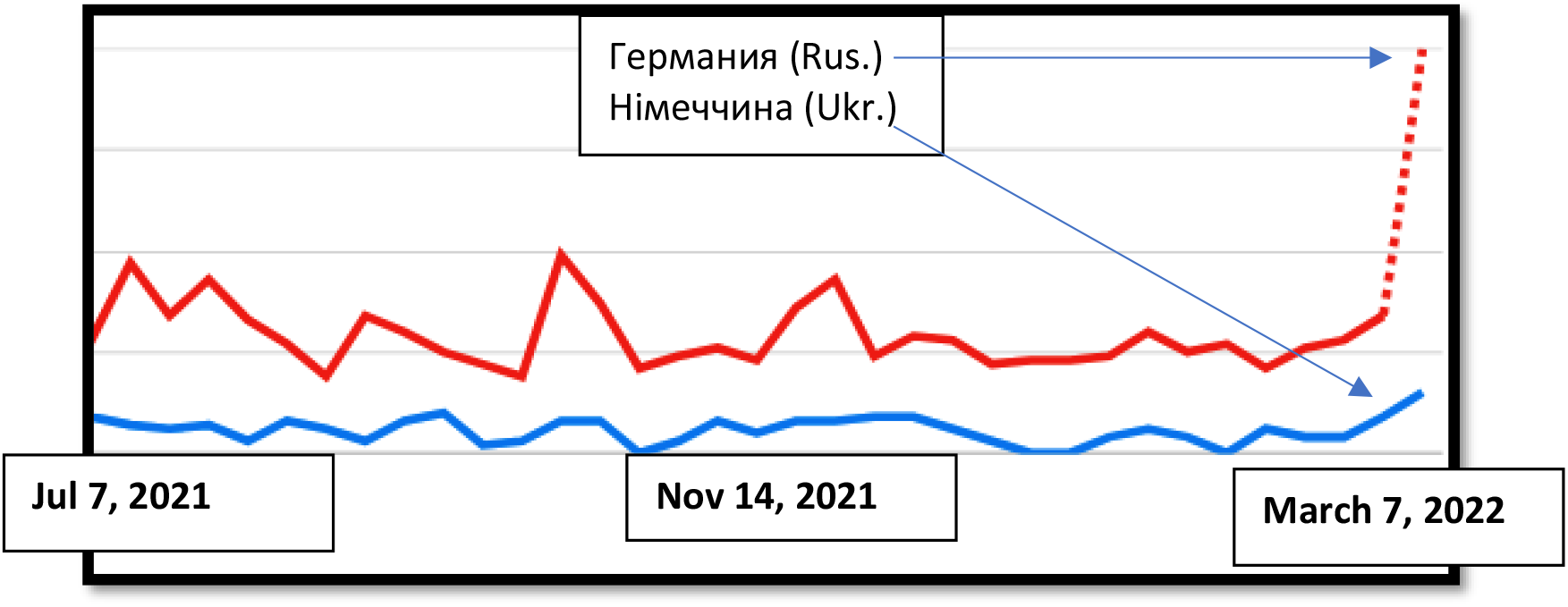
The search term “Німеччина, Германия” (Germany) in Poland (from July 11, 2021, to Macrh 08, 2022)

**Figure 11.**
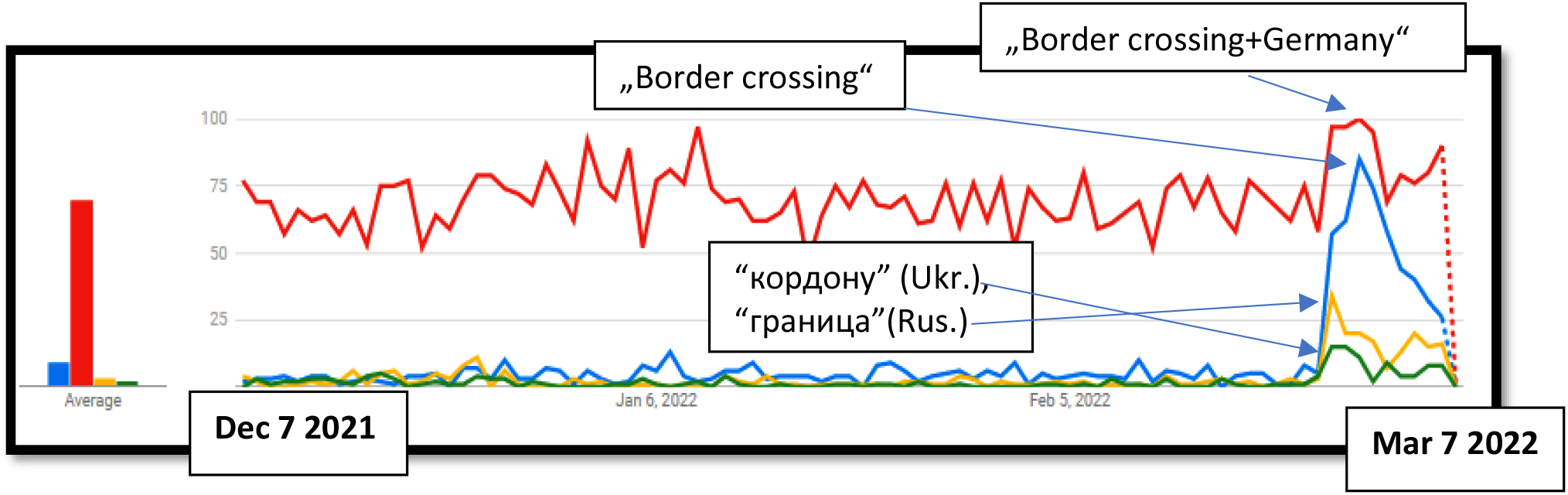
A most common Internet search in Poland: „Border crossing+Germany“, “border crossing”, “кордону” (Ukr. border), “граница” (Russ. border) (December 7, 2021, to March 7, 2022)

**Figure 12.**
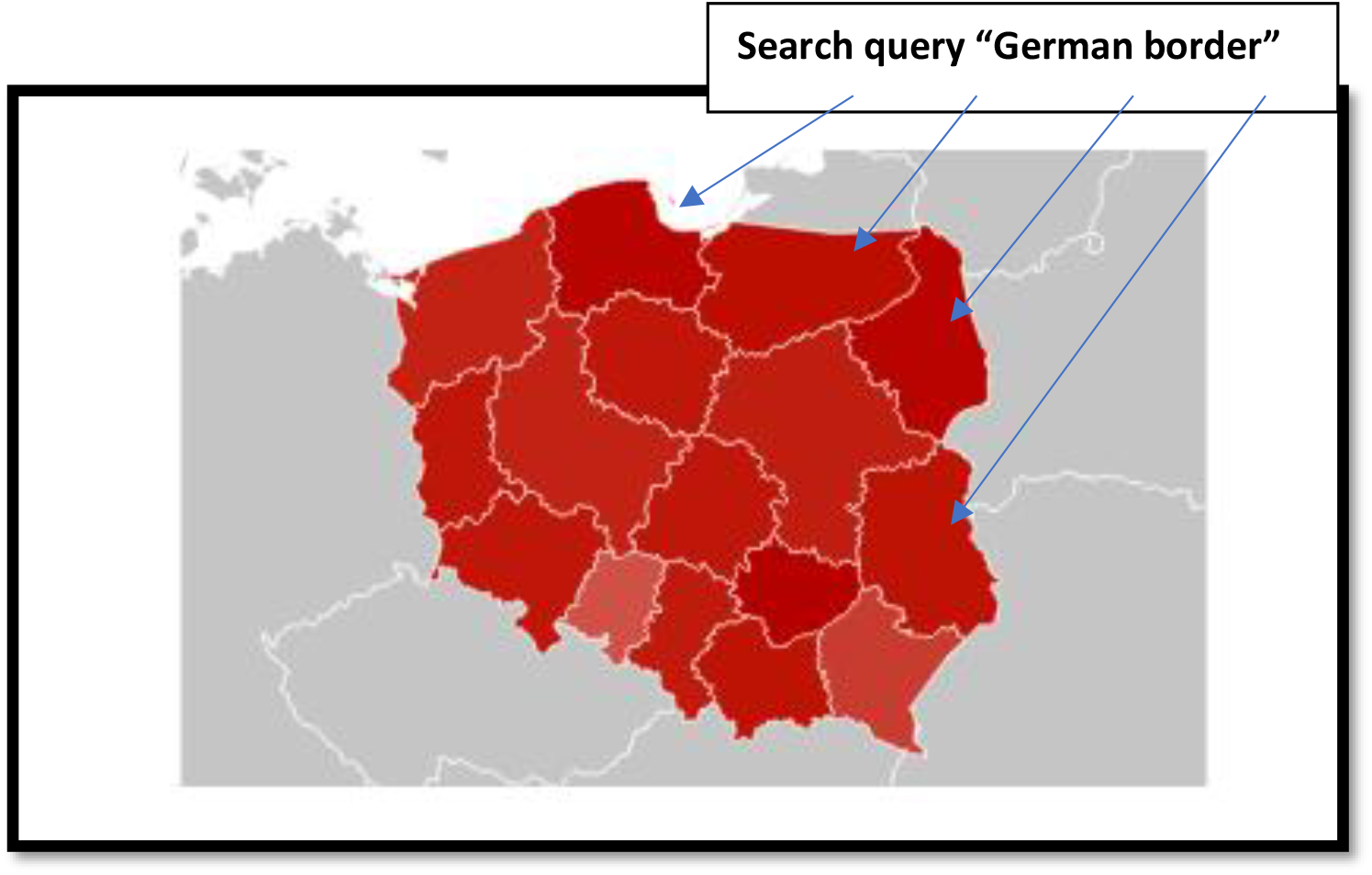
Regions in Poland where interest for “the German border” is growing (March 6 2022)

**Figure 13.**
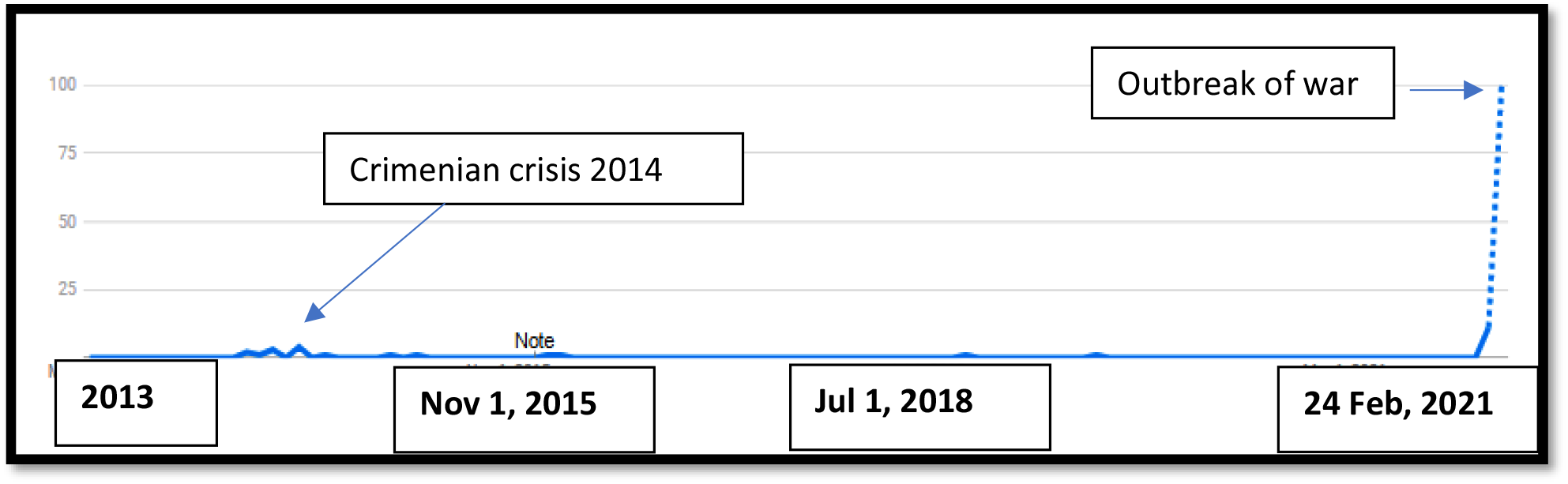
Comparison of the search queries “Германия принимает беженцев из Украины” (Germany accepts refugees from Ukraine) during the Annexation of Crimea in 2014 with the current crisis in 2022.

In further proceedings to standardise the data, we requested the data from February 1 to March 11, 2022, and divided the keyword frequency for the most searched terms “граница” and “кордону” (border) and compared this search index with official statistics from UNHCR to prove the signification of results.

**Table 3.** Refugees from Ukraine in Poland, Hungary and West European countries.

| Datum | Poland | Hungary | Other European countries (West) |
| --- | --- | --- | --- |
| 11. Mar 2022 | 1,524,903 | 225,046 | 282,497 |
| 08. Mar 2022 | 1,204,403 | 191,348 | 210,239 |
| 04. Mar 2022 | 756,303 | 157,004 | 133,876 |
| 28. Feb 2022 | 280.000 | 85.000 | 34,600 |
| **Total: 2,504,893 (Data as 11 Mar 2022)** |  |  |  |

Source: UNHCR, https://data2.unhcr.org/en/situations/ukraine and UNHCR, Ukraine Situation Regional Refugee Response Plan, https://data2.unhcr.org/en/situations/ukraine, authors creation

Figure 14 shows that the increase in Google search for the query “граница” (border) in Russian correlates with the increase of externally displaced persons from Ukraine to Poland (Feb 24 – Mar 11, 2022).

**Figure 14.**
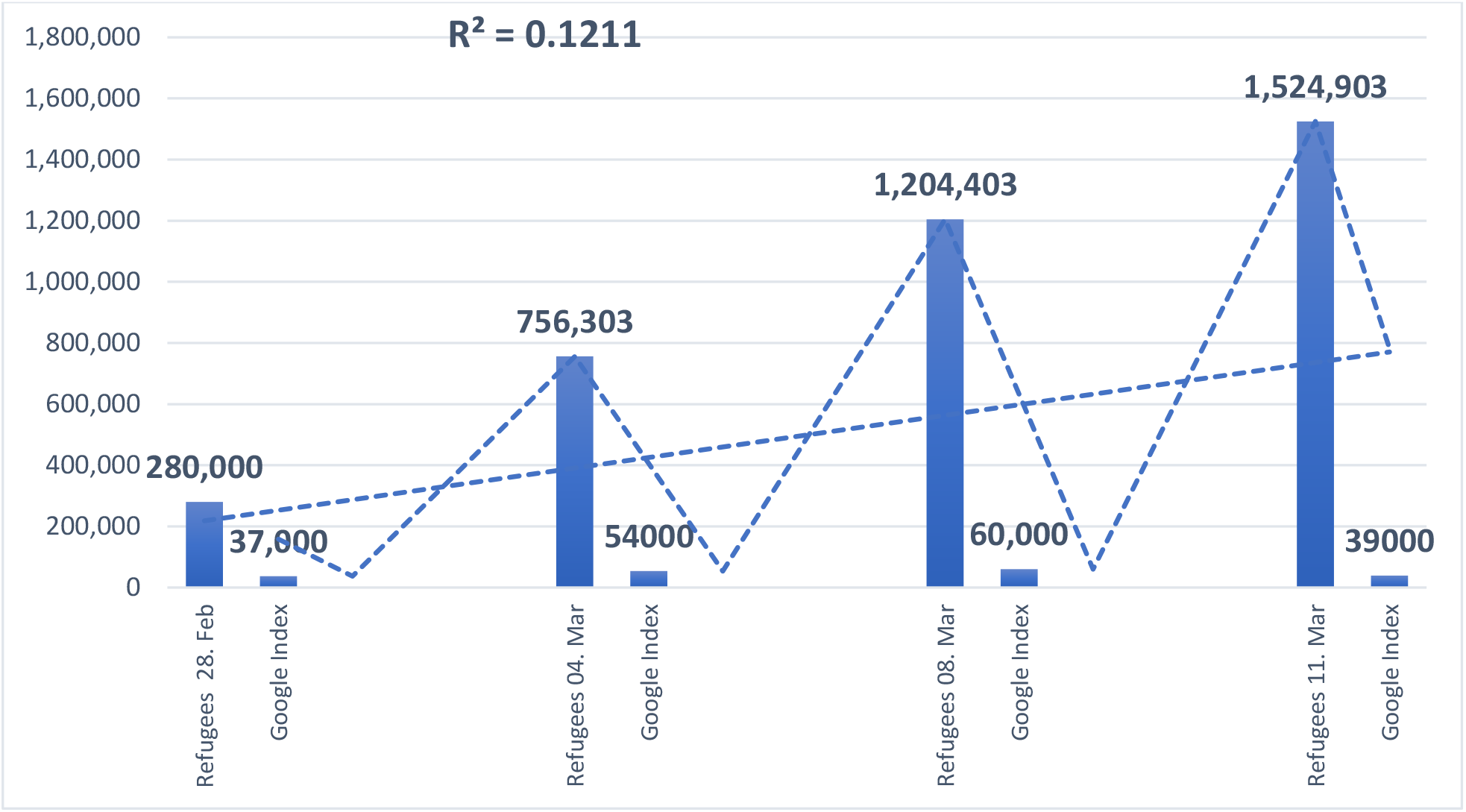
Correlation between Google search index for query “граница” (border) in Russian and the UNHCR statistics for externally displaced persons from Ukraine to Poland (24.02. – 11.03.2022)

**Figure 15.**
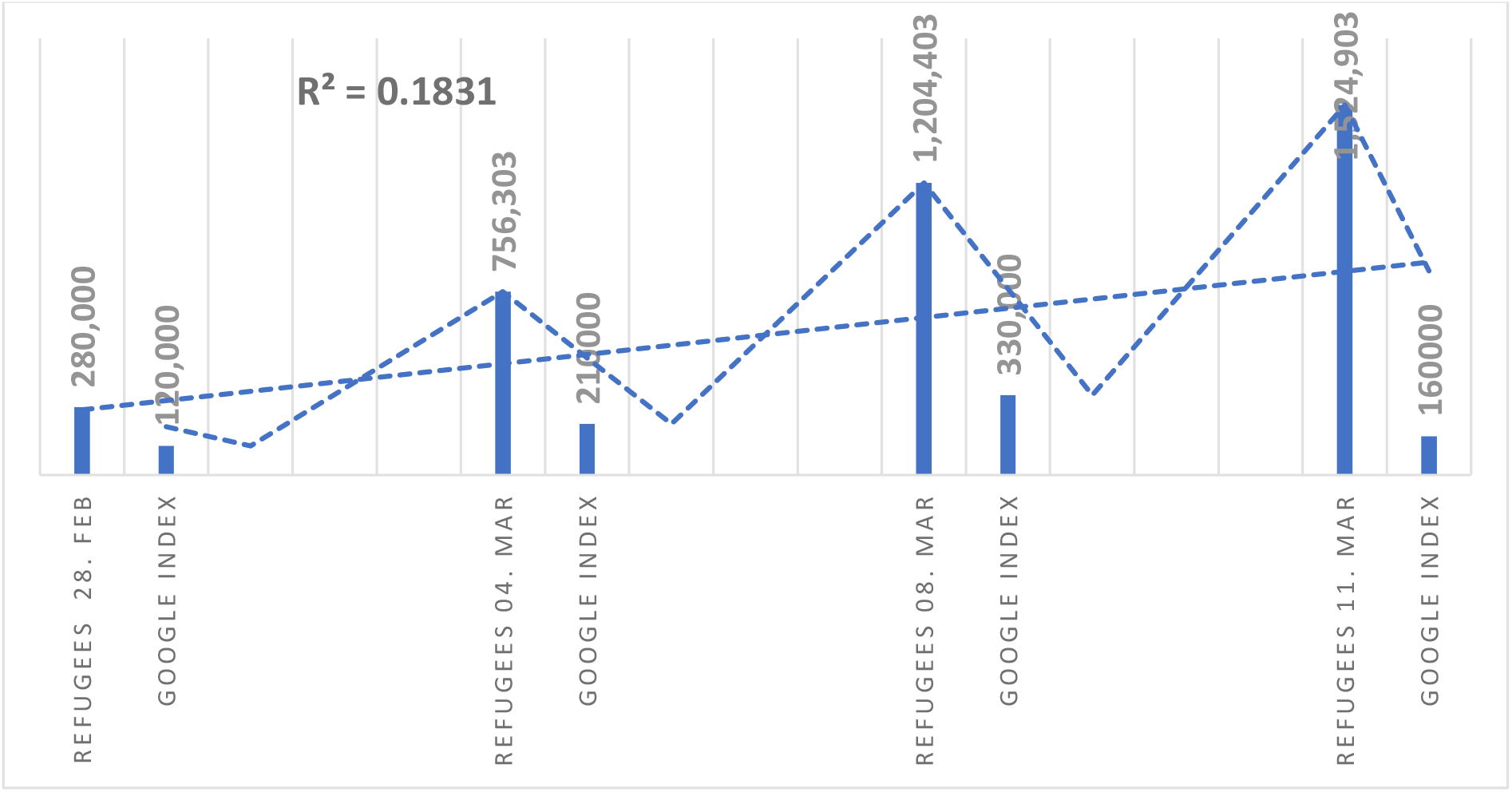
Correlation between Google search index for query “кордону” (border) in Ukrainian and the UNHCR statistics for externally displaced persons from Ukraine to Poland (Feb 24 – Mar 11, 2022)

Figure 14 shows that there is a positive correlation as in the case of Google searches in Russian. The increase in Google search for the query “кордону” (border) in Ukrainian correlates with the rise of externally displaced persons from Ukraine to Poland (February 24 – March 11 2022).

Our previous studies^87^ have shown that in opposite when the Google search decrease, there will be a decrease in externally displaced persons from Ukraine. There is a positive linear association between the Google index and data from official statistics, UNHCR, enabling reliable estimates and forecasting for the future.

### Social media as a source of refuge flows data

Since UNHCR does not yet have complete data on the demographic structure of refugees, and GT does not provide any demographic data on the sample structure to fill this gap, we used two other alternative Big Data sources: Facebook and Instagram. When withdrawing Big Data records from Instagram and Facebook to determine the demographic structure of refugees, we used their insights tools.

The results show that the share of women in the refugee population from Ukraine is 61.3% in Poland and 60.6% in Germany. When we set the filter to limit the specific interests that only Ukrainians in Poland could have and keep the Ukrainian language option, we get a more accurate picture showing an estimate of 246,300 people using FB and Instagram in Ukrainian and a demographic structure of 65.1% of women. The age group most represented is between 24 and 45 years (data for children are not available).

It is especially interesting to note that in the period in which we monitored this data, we noticed every day that the number of users of FB and Instagram in Ukrainian in Poland grew by 7,000 to 10,000, while in Germany, it grew by 1,000 to 2,000 new users. Collecting data from FB and Instagram is a particular problem for the researcher as he has to re-collect data every day and keep his own archive because FB and Instagram do not provide retroactive insight for past periods as GT It is also necessary to take into account the overall social context and the fact that even before the war, a large number of Ukrainians emigrated to Poland. 1,4 million Ukrainians lived in Poland before the war broke out (most fled after the Crimea crisis). It is important to note also that many Ukrainians use FB and Instagram in Russian (but also in English). As official data have not yet been released, insights that we gained from Facebook and Instagram that most refugees are in Warsaw, Breslau, and Krakow we could not yet verify.

Twitter is also an insightful tool that, especially with its conversation tracking option, enables monitoring the increase in interactions and the growth of interest in topics related to migration.^88^ Geo-referenced social media posts and micro-blogs, particularly geotagged Tweets, have been a popular source to track forced displacement streams.^89^ The basic idea is to use forced displaced people’s social media activities as a tracking device for their movements or new place of residence.^90^ A significant limitation is that only around 3% of all Tweets are geotagged.^91^ Tweets provide information on the location, the user’s nationality, birthplace, and the user’s language chosen when generating their account.

Finally, YouTube insights have also proven to be a valuable alternative database. In our analysis, we used YouTube topics related to life and work in Germany and Poland since the outbreak of war in Ukraine, which as well as keyword searches indicated an increase in interest.

## Discussion

All tested migration-related search queries, which show an indication about emigration planning, shows a positive linear association between Google index and data from official statistics, UNHCR; R^2^ = 0.1211 for searches in Russian and R^2^ = 0.1831 for searches in Ukrainian.

The increase in Google search is correlated with the rise in the number of refugees in the EU. In contrast, according to our previous studies, the decrease in Google search will show the decrease in externally displaced persons.

Out testing shows the increase in Internet searches for all neighbouring countries in Russian, Ukrainian and English; all three languages shows that the interest for Poland is the highest. It should be emphasised that when refugees arrive in nearby countries, the search for terms related to *Germany*, such as “crossing the border + Germany”, etc., is proliferating. This result confirms our hypothesis that up to one-third of all refugees will cross into Germany.

Regarding the estimated number of expected refugees, we compared the Crimean crisis in 2014 and the current crisis. During the peak of the Crimean crisis in 2014, the search index for query “Германия принимает беженцев из Украины” (Germany accepts refugees from Ukraine) was 4, and in 2022 it is 12, that is, the interest in fleeing the country is now three times higher. Therefore, according to Big Data insights, it is to expect 5,4 Million refugees.

Since UNHCR does not yet have complete data on the demographic structure of refugees, and GT does not provide any demographic data on the sample structure to fill this gap, we used two other alternative Big Data sources: Facebook and Instagram. Although these insights could not answer the question about the demographic structure of refugees, they provided some valuable insights that could be a very useful alternative source of data with further improvement of the method.

The presented method regarding GT contributes in a way that proves the feasibility of predicting further refugees wave from Ukraine, which allows reliable forecasts for the future and helps prepare humanitarian aid and civil infrastructure. This procedure also presents a new methodological approach to how data obtained through GT can be standardised for comparison with official UNHCR databases. The insights are particularly relevant for EU policymakers and can help governments to design appropriate strategies and prepare and better respond to this crisis in the future. Namely, even if the war ends quickly, Croatia’s previous experience with the war shows that the crisis in Ukraine will continue for many years to come, and it is to be expected that the abolition of military service will lead to the emigration of large numbers of men from the country.

## Conclusion

This study was created as a result of the need to predict migration flows of refugees from Ukraine to the EU and to prepare the humanitarian infrastructure. The usefulness and the main advantage of this approach is the timely identification of external migrations from Ukraine, which can be used to model projections and predict future trends. We used linear regression to measure the correlation between the number of searches (x) and the number of moves (y) evidenced by the official UNHCR statistics. There was a clear correlation between the Google Trends index, and refugee flows from Ukraine.

Analysis of digital traces showed that refugees most often searched for the term “border” in Ukrainian, Russian and English. The data generated by our approach correlates with official UNHCR data regarding the direction of refugee flows from Ukraine, but not with an estimated number. Our insights show that 27% more refugees are expected than the UN shows.

According to geolocations, the tested crosschecks of migration-related searches correspond to the official UNHCR data. Big (Crisis) Data show that the main regions of emigration from Ukraine are the provinces directly affected by the war - as expected.

Our study shows that refugees will not necessarily stay in the countries of first immigration and that almost one third will continue their journey to Germany. According to the Big (Crisis) Data approach, in the next 12 months, Germany can expect 1,5 Million Ukrainian refugees. Since UNHCR does not yet have complete data on the demographic structure of refugees, and GT does not provide any demographic data on the sample structure to fill this gap, we used two other alternative Big Data sources: Facebook and Instagram.

Despite the many advantages of Big (Crisis) Data, there are still significant open methodological issues. At the same time, more and more studies argue that samples obtained from Big Data do not significantly differ from samples obtained from more traditional recruitment and sampling techniques.^92^ Although these open-ended issues pose severe challenges for making precise estimates, statistics offer various tools to deal with imperfect data.^93^ Despite its limitations, we believe that the model presented here can contribute to the migration and refuge studies, especially now in the Ukrainian refugee crisis and that Big Data hold huge potential for humanitarian organisations and governments to improve their early warning systems.

## Data Availability

All data produced in the present work are contained in the manuscript

## Funding

This research received no external funding.

## Compliance with ethical standards

In this work, we use only anonymous, aggregate data. All data are collected following the applicable GDPR and ethical principles of personal data handling. This project’s database retains no information about the identity, IP address, or specific physical location of any user.

## Conflicts of Interest

The author declares no conflict of interest.

## Footnotes

1 UNHCR, https://data2.unhcr.org/en/situations/Ukraine (04.03.2022)

2 Ibid.

3 UNHCR Global Data Service (2021). Big (Crisis) Data for Predictive Models. A Literature Review, p 11.

4 Ibid.

5 Spyratos, S., M. Vespe, F. Natale, I. Weber, E. Zagheni, and M. Rango (2018). Migration Data using Social Media. A European Perspective. JRC Technical Reports European Commission. Luxembourg: Publications Office of the European Union.

6 Wanner, P. (2020). “How Well CanWeEstimate Immigration Trends Using Google Data?” Quality and Quantity 55: 1181–202.

7 Britannica.com, Ukraine: History – Britannica Online Encyclopedia (06.03.2022)

8 Bergtora Sandvik, K. and Garnier A. (4 Mar 2022). Forced displacement from Ukraine: notes on humanitarian protection and durable solutions, https://reliefweb.int/report/ukraine/forced-displacement-ukraine-notes-humanitarian-protection-and-durable-solutions (07.03.2022)

9 Ibid.

10 Ibid.

11 See: Bergtora Sandvik, K. and Garnier A. (4 Mar 2022). Forced displacement from Ukraine …

12 Kuznetsova, I. and Mikheieva, O. (2020) Forced displacement from Ukraine’s war-torn territories: intersectionality and power geometry. Nationalities Papers 48.4 (2020): 690-706

13 Charron, A. (2020). Somehow, we cannot accept it’: Drivers of internal displacement from Crimea and the forced/voluntary migration binary. Europe-Asia Studies 72.3 (2020): 432-454.

14 Sereda, V. (2020). Social Distancing’and Hierarchies of Belonging: The Case of Displaced Population from Donbas and Crimea. Europe-Asia Studies 72.3 (2020): 404-431

15 Scrinic, A. (2014). Humanitarian aid and political aims in Eastern Ukraine: Russian involvement and European response. Eastern Journal of European Studies 5.2 (2014): 77-88.

16 UNHCR, Ukraine Situation Regional Refugee Response Plan, https://data2.unhcr.org/en/situations/Ukraine (04.03.2022)

17 UNHCR, Syria RRR, https://data2.unhcr.org/en/situations/Syria (08.03.2022)

18 UNHCR head Filippo Grandi, The Guardian (05.03.2022)

19 UNHCR, https://data2.unhcr.org/en/situations/ukraine (08.03.2022)

20 See: Radelić Z., D Marijan, N. Barić, A. Bing, D. Živić, *Stvaranje hrvatske države i Domovinski rat*, Školska knjiga Zagreb 2006.

21 Jurić, T. *“Gastarbeiter Millennials”. Exploring the past, present and future of migration from Southeast Europe to Germany and Austria with approaches to classical, historical and digital demography*. Hamburg 2021, Verlag Dr. Kovač. p 33.

22 Kristin Bergtora Sandvik and Adèle Garnier, (4 Mar 2022). Forced displacement from Ukraine: notes on humanitarian protection and durable solutions, https://reliefweb.int/report/ukraine/forced-displacement-ukraine-notes-humanitarian-protection-and-durable-solutions (07.03.2022)

23 Visitukraine, https://visitukraine.today/blog/154/refugees-from-ukraine-received-the-right-to-live-in-the-eu-for-3-years (11.03.2022)

24 Zuwanderungsbedarf aus Drittstaaten in Deutschland bis 2050. Bertelsmann Stiftung. 2015. URL: https://www.bertelsmann-stiftung.de/fileadmin/files/BSt/Publikationen/GrauePublikationen/ (24.02.2022)

25 Arbeitsagentur (2020). Gemeldete Arbeitsstellen nach Berufen (Engpassanalyse), https://statistik.arbeitsagentur.de/Statistikdaten/ (24.02.2022)

26 Bertelsmann Stiftung (2015). Zuwanderungsbedarf aus Drittstaaten in Deutschland bis 2050.

27 Ibid.

28 Deutsche Welle, Ukrajinske izbjeglice kao konkurencija useljenicima sa zapadnog Balkana?, https://www.dw.com/bs/ukrajinske-izbjeglice-kao-konkurencija-useljenicima-sa-zapadnog-balkana/a-61045150

29 Abcnews (2022). https://abcnews.go.com/International/europes-unified-ukrainian-refugees-exposes-double-standard-nonwhite/story?id=83251970

30 Maissaa Almustafa (2021). Reframing refugee crisis: A “European crisis of migration” or a “crisis of protection”?, https://doi.org/10.1177/2399654421989705

31 BMI (2015), https://www.bmi.bund.de/SharedDocs/kurzmeldungen/EN/2015/09/measures-on-asylum-and-refugee-policy.html (25.02.2022)

32 Berkay M. (2020). Sharing the Burden: Revisiting the EU-Turkey Migration Deal, https://www.crisisgroup.org/europe-central-asia/western-europemediterranean/turkey/sharing-burden-revisiting-eu-turkey-migration-deal (25.02.2022)

33 Jurić, T. (2022). Forecasting Migration and Integration Trends Using Digital Demography – A Case Study of Emigration Flows from Croatia to Austria and Germany, Comparative Southeast European Studies, 1-28, https://doi.org/10.1515/soeu-2021-0090

34 e.g. Böhme, M.H., Gröger, A., & Stöhr, T. (2020). Searching for a better life: Predicting international migration with online search keywords. Journal of Development Economics, 142.

35 Wladyka 2017).

36 Jurić, T. (2022). Forecasting Migration and Integration Trends …, https://doi.org/10.1515/soeu-2021-0090

37 UNHCR Blog (Jan 6, 2022). Predicting refugee flows with big data: a new opportunity or a pipe dream? by Andrea Pellandra and Geraldine Henningsen, https://www.unhcr.org/blogs/predicting-refugee-flows-with-big-data-a-new-opportunity-or-a-pipe-dream/ (10.03.2022)

38 Ibáñez Sales, M. (2021). Big data at the crossroads: seizing the potential of Big data to guide the future of EU migration policy. Euromesco Policy brief n. 116, p. 2.

39 Jurić, T. (2021). Medical Brain Drain From Southeastern Europe: Using Digital Demography to Forecast Health Worker Emigration, JMIRx Med 2021, vol. 2, iss. 4, e30831, doi: 10.2196/30831

40 Wang R, Wang W, daSilva A, Huckins JF, Kelley WM, Heatherton TF, et al. (2018) Tracking depression dynamics in college students using mobile phone and wearable sensing, Article No.: 43 https://doi.org/10.1145/3191775

41 Jurić, T. *“Gastarbeiter Millennials”. Exploring the past, present and future of migration from Southeast Europe to Germany and Austria with approaches to classical, historical and digital demography*. Hamburg 2021, Verlag Dr. Kovač. p 98.

42 Cesare N, Lee H, McCormick T, Spiro E, Zagheni E. Promises and Pitfalls of Using Digital Traces for Demographic Research, 2018. DOI:10.1007/s13524-018-0715-2

43 Jurić T. (2021). Primjena analitičkih alata aplikacija YouTube, Google Photo i Google Web u predviđanju dolazaka turista u Republiku Hrvatsku s osvrtom na izazove zdravstvene sigurnosti, Medix, 147/148. URL: https://www.medix.hr/index.php?p=pdf&pdf=google-trends-kao-metoda-za-rano-detektiranje-pojave-novih-slucajeva-covid-a-19 (28.02.2022)

44 Choi H, Varian H. (2012). Predicting the present with google trends. Econ Record 88(s1), 2–9 (2012). https://doi.org/10.1111/j.1475-4932.2012.00809.x

45 Önder I. (2017). Forecasting Tourism Demand with Google Trends: Accuracy Comparison of Countries vs. Cities, 2017. https://doi.org/10.1002/jtr.2137

46 Agrawal A., Sahdev R.; Davoudi H.; Khonsari F.; An A.; McGrath S. (2016). Detecting the Magnitude of Events from News Articles, https://ieeexplore.ieee.org/document/7817051

47 Singh et al. (2019). KDD ‘19: Proceedings of the 25th ACM SIGKDD International Conference on Knowledge Discovery & Data Mining, p. 1975–1983, https://doi.org/10.1145/3292500.3330774

48 UNHCR Global Data Service (2021). Big (Crisis) Data for Predictive Models. A Literature Review, p 11.

49 Jurić, T. (2022). Forecasting Migration and Integration Trends Using Digital Demography – A Case Study of Emigration Flows from Croatia to Austria and Germany, Comparative Southeast European Studies, 1-28, https://doi.org/10.1515/soeu-2021-0090

50 United Nations (2014) The Data Revolution for Human Development. http://hdr.undp.org/en/content/data-revolution-human-development (accessed 28.02.2022); United Nations (2019). Report of the Global Working Group on Big Data for Official Statistics. The Economic Social Council of the United Nations, UN Global Working Group on Big Data. https://unstats.un.org/unsd/statcom/51st-session/documents/2020-24-BigData-E.pdf (27.02 2022).

51 Zagheni, E., I. Weber, and K. Gummadi. 2017. “Leveraging Facebook’s Advertising Platform to Monitor Stocks of Migrants.” Population and Development Review 43 (4): 721–34; Zagheni, E., and I. Weber. 2015. “Demographic Research with Non-Representative Internet Data.” International Journal of Manpower 36 (1): 13–25.

52 e. g. Dubois et al. 2018; Hawelka et al. 2014; State et al. 2014, Jurić 2021, 2022.

53 Jurić, T. (2021). Medical Brain Drain From Southeastern Europe: Using Digital Demography to Forecast Health Worker Emigration, JMIRx Med 2021, vol. 2, iss. 4, e30831, doi: 10.2196/30831

54 Spyratos, S., M. Vespe, F. Natale, I. Weber, E. Zagheni, and M. Rango. 2018. Migration Data using Social Media. A European Perspective. JRC Technical Reports European Commission. Luxembourg: Publications Office of the European Union.

55 Jurić, T. (2022). Forecasting Migration and Integration Trends …, https://doi.org/10.1515/soeu-2021-0090

56 Gabrilovich, E. 2020. Using Symptoms Search Trends to Inform COVID-19 Research. Google Health. https://blog.google/technology/health/using-symptoms-search-trends-inform-COVID-19-research (27.02.2022).

57 Wanner P. (2020). How well can we estimate immigration trends using Google data?, Quality & Quantity, 2020. https://doi.org/10.1007/s11135-020-01047-w

58 Connor, P. (2017). The digital footprint of Europe’s refugees. Methodology. Pew Research Center. 2017. URL: https://www.pewglobal.org/2017/06/08/online-searches-eu-refugees-methodology/ [accessed 20.07.2021]

59 Jurić T. (2022). Facebook i Google kao empirijska osnova za razvoj metode digitalnog praćenja vanjskih migracija hrvatskih građana, Ekonomski pregled (forthcoming)

60 Jurić, T. (2021). Using Digital Demography to Forecast Health Worker Emigration, JMIRx Med 2021, doi: 10.2196/30831

61 Internet users distributions in the world - 2021. Internet World Stats. https://www.internetworldstats.com/stats.htm (02.03.2022)

62 Ibáñez Sales, M. (2021). Big data at the crossroads: seizing the potential of Big data to guide the future of EU migration policy. Euromesco Policy brief n. 116, p. 2.

63 See: Connor, P. The digital footprint of Europe’s refugees. Methodology. Pew Research Center. 2017. URL: https://www.pewglobal.org/2017/06/08/online-searches-eu-refugees-methodology/ [accessed 20.07.2021]

64 Google Trends. https://trends.google.com/trends/?geo=HR

65 Jurić, T. (2021). Google Trends as a Method to Predict New COVID-19 Cases and Socio-Psychological Consequences of the Pandemic, Athens Journal of Mediterranean Studies 2021, 7: 1-25, https://www.athensjournals.gr/mediterranean/2021-4210-AJMS-Juric-05.pdf

66 Jurić, T. (2022). Forecasting Migration and Integration Trends Using Digital Demography – A Case Study of Emigration Flows from Croatia to Austria and Germany, Comparative Southeast European Studies, 1-28, https://doi.org/10.1515/soeu-2021-0090

67 Jurić, T. *“Gastarbeiter Millennials”. Exploring the past, present and future of migration from Southeast Europe to Germany and Austria with approaches to classical, historical and digital demography*. Hamburg 2021, Verlag Dr. Kovač. p 260.

68 Jurić, T. (2021). Medical Brain Drain From Southeastern Europe: Using Digital Demography to Forecast Health Worker Emigration, JMIRx Med 2021, vol. 2, iss. 4, e30831, doi: 10.2196/30831; Jurić, T. (2022). Forecasting Migration and Integration Trends Using Digital Demography – A Case Study of Emigration Flows from Croatia to Austria and Germany, Comparative Southeast European Studies, 1-28, https://doi.org/10.1515/soeu-2021-0090.

69 See: Jurić, T. *“Gastarbeiter Millennials”. Exploring the past, present and future of migration from Southeast Europe … 2021*.

70 Curry T., Croitoru A., Crooks, A., Stefanidis A. (2018). Exodus 2.0: crowdsourcing geographical and social trails of mass migration. Journal of Geographical Systems.

71 See further explanations byWanner, P. 2020. “How Well CanWeEstimate Immigration Trends Using Google Data?” Quality and Quantity 55: 1181–202.; and Wilde, J., W. Chen, and S. Lohmann. 2020. “COVID-19 and the Future of US Fertility: What Can We Learn from Google?” IZA Discussion Papers 13776. www.iza.org/publications/dp/13776/ covid-19-and-the-future-of-us-fertility-what-can-we-learn-from-google and

72 UNHCR Blog (Jan 6, 2022). Predicting refugee flows with big data: a new opportunity or a pipe dream? by Andrea Pellandra and Geraldine Henningsen, https://www.unhcr.org/blogs/predicting-refugee-flows-with-big-data-a-new-opportunity-or-a-pipe-dream/ (10.03.2022)

73 UNHCR Global Data Service (2021). Big (Crisis) Data for Predictive Models. A Literature Review, p 19

74 Ibid, p 7.

75 Wladyka D. (2017). Queries to Google Search as predictors of migration flows from Latin America to Spain. J Popul Soc Stud 2017 Oct 1;25(4):312-327. [doi: 10.25133/jpssv25n4.002]

76 EUROPOL (2018). Two years of EMSC: Activity report January 2017 – January 2018. European Migration Smuggling Centre, EUROPOL. Retrieved from: https://www.europol.europa.eu/cms/sites/default/files/documents/two_years_of_emsc_report.pdf (10.03.2022)

77 UNHCR (2017). From a refugee perspective: Discourse of Arabic speaking and Afghan refugees and migrants on social media from March to December 2016, https://www.unhcr.org/publications/brochures/5909af4d4/from-arefugee-perspective.html https://reliefweb.int/sites/reliefweb.int/files/resources/58018.pdf (10.03.2022)

78 Jurić, T. (2022). Forecasting Migration and Integration Trends Using Digital Demography – A Case Study of Emigration Flows from Croatia to Austria and Germany, Comparative Southeast European Studies, 1-28, https://doi.org/10.1515/soeu-2021-0090

79 Wladyka D. (2017). Queries to Google Search as predictors of migration flows from Latin America to Spain. J Popul Soc Stud 2017 Oct 1;25(4):312-327. [doi: 10.25133/jpssv25n4.002]

80 Jurić, T. *“Gastarbeiter Millennials”. Exploring the past, present and future of migration from Southeast Europe to Germany and Austria with approaches to classical, historical and digital demography*. Hamburg 2021, Verlag Dr. Kovač.

81 UNHCR Global Data Service (2021). Big (Crisis) Data for Predictive Models. A Literature Review, p 11.

82 Matías Ibáñez Sales (2021). Big data at the crossroads: seizing the potential of Big data to guide the future of EU migration policy. Euromesco Policy brief n. 116, p. 8.

83 Bircan, T., & Korkmaz, E.E. (2021). Big data for whose sake? Governing migration through artificial intelligence. Humanities Social Sciences Communications, 8, https://www.nature.com/articles/s41599-021-00910-x

84 BMI, https://www.bmi.bund.de/SharedDocs/faqs/EN/topics/ministry/ukraine-war-eng/faq-ukraine-artikel.html

85 UNHCR, Internally Displaced Persons (IDP), https://www.unhcr.org/ua/en/internally-displaced-persons

86 Mediendiesnt Integration. Krieg in der Ukraine: Wie ist die Flüchtlings-Situation?, Franck Düvell, Universität Osnabrück, https://mediendienst-integration.de/artikel/krieg-in-der-ukraine-wie-ist-die-fluechtlings-situation.html (07.03.2022)

87 Jurić, T. (2021). Medical Brain Drain From Southeastern Europe: Using Digital Demography to Forecast Health Worker Emigration, JMIRx Med 2021, vol. 2, iss. 4, e30831, doi: 10.2196/30831; Jurić, T. (2022). Forecasting Migration and Integration Trends Using Digital Demography – A Case Study of Emigration Flows from Croatia to Austria and Germany, Comparative Southeast European Studies, 1-28, https://doi.org/10.1515/soeu-2021-0090

88 Zagheni E., Garimella V.R.K., Weber I. and State B. (2014). Inferring International and Internal Migration Patterns from Twitter Data. 439-444.

89 UNHCR Global Data Service (2021). Big (Crisis) Data for Predictive Models. A Literature Review, p 19

90 Alina Sirbu, Gennady Andrienko, Natalia Andrienko, Chiara Boldrini, Marco Conti, Fosca Giannotti, Riccardo Guidotti, Simone Bertoli, Jisuand Kim, Cristina Ioana Muntean, Luca Pappalardo, Andrea Passarella, Dino Pedreschi, Laura Pollacci, FrancescaPratesi, and Rajesh Sharma. Human migration: the big data perspective. International Journal of Data Science and Analytics, March 2020.

91 Alessandra Righi. Assessing migration through social media: a review. Mathematical Population Studies, 2019.

92 Kalimeri, K., A. Bonanomi, M. Beiro, A. Rosina, and C. Cattuto (2020). Traditional versus Facebook-Based Surveys: Evaluation of Biases in Self-Reported Demographic and Psychometric Information. Demographic Research 42: 133–48.

93 Cesare N, Lee H, McCormick T, Spiro E, Zagheni E. (2018). Promises and pitfalls of using digital traces for demographic research. Demography 2018 Oct;55(5):1979-1999, doi: 10.1007/s13524-018-0715-2, Medline: 30276667

## Notes

### Competing Interest Statement

The authors have declared no competing interest.

### Funding Statement

This study did not receive any funding.

### Author Declarations

Ethics committee/IRB of Catholic University of Croatia gave ethical approval for this work

